# Symptom presentation among SARS-CoV-2 positive cases and the impact of COVID-19 vaccination; three prospective household cohorts

**DOI:** 10.1101/2022.08.19.22278985

**Authors:** Ilse Westerhof, Marieke de Hoog, Margareta Ieven, Christine Lammens, Janko van Beek, Ganna Rozhnova, Dirk Eggink, Sjoerd Euser, Joanne Wildenbeest, Liesbeth Duijts, Marlies van Houten, Herman Goossens, Carlo Giaquinto, Patricia Bruijning-Verhagen

**Affiliations:** Julius Centre for Health Sciences and Primary Care, Department of Epidemiology, University Medical Centre Utrecht, Utrecht, The Netherlands; Laboratory of Medical Microbiology, Vaccine & Infectious Disease Institute, University of Antwerp, Antwerp, Belgium; Department of Viroscience, Erasmus University Medical Center Rotterdam, Rotterdam, The Netherlands; BioISI—Biosystems & Integrative Sciences Institute, Faculdade de Ciências, Universidade de Lisboa, Lisboa, Portugal; Centre for Infectious Disease Control, WHO COVID-19 Reference Laboratory, National Institute for Public Health and the Environment (RIVM), Bilthoven, The Netherlands; Streeklaboratorium voor de Volksgezondheid Kennemerland, Department of Epidemiology, Kennemerland, The Netherlands; Department of Pediatric Infectious Diseases and Immunology, Wilhelmina Children’s Hospital University Medical Center Utrecht, Utrecht, The Netherlands; Erasmus MC - Sophia Children’s Hospital, Erasmus University Medical Center, Department of Pediatrics, Rotterdam, The Netherlands; Spaarne Gasthuis Academy, Spaarne Gasthuis, Hoofddorp, The Netherlands; Spaarne Gasthuis, department of Pediatrics, Haarlem and Hoofddorp, The Netherlands; Department of Women and Child Health, University of Padova, Padova, Italy

## Abstract

**OBJECTIVE:** Using data from European prospective household studies, we systematically compared the symptom burden of the wild-type and Alpha variant infected individuals versus the Omicron BA.1 and BA.2 infected individuals across paediatric and adult age-groups. In addition, we measured the impact of COVID-19 vaccination on the Omicron symptom burden.

**METHODS:** The household transmission studies were conducted during the wild-type and Alpha period (April 2020 to April 2021) and the early Omicron BA.1 and BA.2 dominant period (January to March 2022). All three studies used similar protocols. Households were prospectively followed from detection of the first SARS-CoV-2 index case until at least day 21 including (repeated) PCR testing, paired serology and daily symptom reporting for all household members. To avoid possible index-case ascertainment bias, we restricted analyses to secondary household cases. Age-stratified SARS-CoV-2 symptom burden was compared for wild-type/Alpha versus Omicron infections and for primary versus primary plus booster series vaccinated adult cases.

**FINDINGS:** In total 216 secondary cases from wild-type/Alpha, and 130 from the Omicron period were included. Unvaccinated children <12 years experienced more symptoms and higher maximum and cumulative severity scores during the Omicron compared to the wild-type/Alpha period (p=0.004, p=0.011 and p=0.075, respectively). In adults, disease duration and maximum and cumulative severity scores were reduced during the Omicron period. Adjusted for age, gender and prior immunity Omicron was associated with lower odds for loss of smell or taste (Odds Ratio [OR]: 0.14; 95%CI 0.03-0.50), and higher, but non-significant odds for upper respiratory symptoms, fever and fatigue (ORs varying between 1.85-2.23). Comparing primary versus primary plus booster vaccinated adult cases during the Omicron period no differences were observed in disease severity or duration (p≥0.12).

**INTERPRETATION:** In children, the Omicron variant causes higher symptom burden compared to the wild-type/Alpha. Adults experienced a lower symptom burden possibly due to prior vaccination. A shift in most frequently reported symptoms occurred with a marked reduction in loss of smell or taste during the Omicron period. An additional effect of booster vaccination on symptom severity in infected adults compared to primary series only, could not be demonstrated.

## Introduction

Since the onset of the Coronavirus Disease 2019 (COVID-19) pandemic in December 2019, several SARS-CoV-2 Variants of Concern (VoC) have emerged^1^. With the emergence of the Omicron VOC (BA.1) and its descendants (BA.2 and further), the pandemic has taken a new turn. Omicron variants are characterized by a high number of mutations compared to the ancestral strain and are associated with immune escape and enhanced ACE-2 binding ^2,3^. In addition, unlike other variants the Omicron variants are less capable of infecting the lower respiratory tract due to a shift in cellular tropism away from TMPRSS2 expressing cells, promoting faster replication in the upper airways but reduced replication in the lungs.^4,5^. As a consequence, the clinical picture of SARS-CoV-2 infection is changing, both in vaccinated and unvaccinated individuals. To date, most studies describing the changing symptomatology of COVID-19 during the Omicron period have focused on cases in hospital.^6,7^ This restricts analyses to more severe cases and therefore provides little understanding on disease evolution in community cases, which represent the overwhelming majority of all cases, and in particular in children, who only rarely require medical care for SARS-CoV-2 infection^8^.

An appropriate setting to study the COVID-19 disease spectrum across paediatric and adult age-groups is within households. We used a dataset from three prospective SARS-CoV-2 household transmission studies that used similar protocols to study age-stratified symptom burden of SARS-CoV2 infections during the wild-type/Alpha dominant period compared to the Omicron BA.1 and BA.2 dominant period, considering differences in vaccination and prior infection status. Our analyses are restricted to secondary household cases to avoid index case ascertainment bias.

## Methods

### Study design and study population

We used data from three prospective household transmission studies conducted in two different SARS-CoV-2 variant periods: the RECOVER household study and CoKids study (Wild-type/Alpha period) and the VERDI-RECOVER household study (Omicron period; Supplement Figure 1-2). The primary aim of the RECOVER household study was to characterize within household transmission of SARS-CoV-2 and the impact of implemented measures within the household to prevent transmission. From April 2020 until April 2021, data from 276 Dutch, Belgium and Swiss households with SARS-CoV-2 were collected. Study design and results on household transmission are published elsewhere.^9^ The Cokids study was set up to determine the susceptibility, transmissibility and disease course of SARS-CoV-2 infection in children. From August 2020 until July 2021, data of 79 SARS-CoV-2 outbreaks of Dutch households with at least one child <18 years of age were collected. Study design and initial results are published elsewhere.^10^ The VERDI-RECOVER household study recruited from January to March 2022 during the period when Omicron BA.1 and BA.2 were dominant. This study aims to estimate household transmission rates in partially vaccinated populations and explores the viral kinetics in Omicron variant infected subjects. During wild-type/Alpha period, vaccination became available for adults (from January 2021 onwards). During the Omicron period, booster vaccinations were available for adults (in the Netherlands from December 2021 onwards) and primary series for adolescents (in the Netherlands from July 2021 onwards). Vaccination for children aged 5 to 11 became available for medical risk groups during mid-January 2022 in the Netherlands.

In all three studies, household outbreaks had started with the identification of an index case followed by repeated sampling and daily symptom monitoring in all household members until the outbreak ended. Enrolment took place within 48 hours following a positive SARS-CoV-2 PCR test of the index case. Households were excluded if one or more household members did not consent to participate. In the VERDI-RECOVER household study, the additional exclusion criterium was SARS-CoV2 positivity in any of the household members in the previous two weeks. Here, these three studies were used to study and compare the COVID-19 disease spectrum across paediatric and adult age-groups between SARS-CoV-2 variants (wild-type, Alpha and Omicron BA.1 and BA.2).

The studies were reviewed and ethically approved by the Medical Ethical Committee Utrecht, The Netherlands (reference number 17-069/M), the Medical Ethical Committee of the Vrije Universiteit university Medical Centre (VUmc), The Netherlands (reference number A2012.901), and the Medical Ethical Committee of Erasmus Medical Centre, The Netherlands (reference number MEC-2020-0609). Written informed consent was obtained from all participating household members and/or their legal representatives.

### Study procedures

Most study procedures were done remotely using self-sampling and an interactive study mobile phone application to accommodate pandemic restrictions on movement, social distancing, and isolation. At baseline, each household member completed a questionnaire including age, comorbidities, recent respiratory complaints, previous infections, and COVID-19 vaccination status. During follow-up, participants daily reported on presence and severity of a set of respiratory and systemic symptoms (see Supplementary Table 1), which was continued until 21 days after last symptom-onset in any household member.

A courier delivered sample kits at home for self-sampling of specimens for viral and serological testing. Self-sampling was supported by live instruction or instruction videos and leaflets, delivered with the sampling material. A telephone helpdesk was available 7 days a week during working hours. In each of the studies, the core protocol included a nose-throat swab (NTS) sample at day 0 for all household members and, if applicable, an additional nose-throat swab from a household member when he/she developed symptoms during follow-up. The core protocol could be extended with additional sample time points and specimens, but this differed between studies (Supplement Table 1). Therefore, those additional results were not used in the current analysis to guarantee similarity in case detection across cohorts. Dried blood spot (DBS) using self-finger-prick were collected at enrolment and end of follow-up.

For data collection on symptoms, questionnaires, and self-sampling we used a custom-made mobile phone application compatible with Apple and Android systems, developed by the UMCU in collaboration with YourResearch Holding BV. The study App contained all study related tasks and questionnaires along with tutorial videos, FAQs, and options to contact the study team. All data entered in the study App were stored in an online secured database. Data were accessible and could be navigated by the study team in real-time by authorized login on the online portal. Daily App-notifications were sent to participants to remind them to complete diary entry and self-sampling when applicable. Study teams received daily reports on participant non-compliance which was followed-up by email, phone, or text-message.

### Laboratory analysis

NTS samples were PCR tested for presence of SARS-CoV-2 and DBS were tested by multiplex protein microarray for antibodies as described previously ^9,10^ at either the Streeklab Haarlem (CoKids cohort), National Institute for Public Health and the Environment (CoKids cohort), Antwerp University (RECOVER and VERDI-RECOVER cohorts), or Erasmus University Medical Center Rotterdam (RECOVER and VERDI-RECOVER cohorts). Details on the PCR and DBS methods used can be found in Supplement 1.

### Definitions

Confirmed SARS-CoV-2 infection was defined as a positive RT-PCR SARS-CoV-2 result or seroconversion defined as SARS-CoV-2 nucleoprotein (NP)-antibody negative at enrolment and positive at the end of follow-up. A secondary household case was defined as a confirmed SARS-CoV-2 infection in a household member not being the index case.

We used the daily symptom data and date of positive test result to define onset and ending of a SARS-CoV-2 episode. An episode started on the day of symptom onset, which had to fall within the seven days prior to, or seven days post first positive test result. An episode ended on the last symptomatic day that was followed by at least two days without any symptoms. SARS-CoV-2 disease severity was categorized into symptomatic disease, pauci-symptomatic, and asymptomatic episodes. Symptomatic disease was defined as: 1) onset of fever OR 2) two consecutive days with one respiratory (cough, sore throat, runny or congested nose, dyspnea) and one systemic symptom (headache, muscle ache, sweats or chills or tiredness) or with at least two respiratory symptoms. Subjects meeting the criteria for symptomatic disease additionally received a daily severity score which consisted of a 5-point Likert scale per reported symptom present, except for fever, which was categorized as <38/38-39/39-40/>40 degrees Celsius. An episode was defined as pauci-symptomatic if symptoms occurred within the specified time-window but remained below the threshold for a symptomatic disease episode, and asymptomatic if no symptoms were reported.

Vaccination status was categorized into unvaccinated, incompletely vaccinated, primary vaccinated, or primary plus booster vaccinated. Primary vaccinated was defined as; two doses of a mRNA vaccine BNT162b2 or mRNA-1273 (Pfizer-BioNtech; Moderna), two doses of the vector-based AZD1222 (AstraZeneca) or a single dose of the vector-based Ad26.COV2.S vaccine (Johnson & Johnson) at least 14 days prior to enrolment. Primary plus booster vaccinated was defined as a third dose of mRNA vaccine, or a second dose if the primary series consisted of the single-dose Ad26.COV2.S vaccine, at least 6 months after completion of the primary series and at least 14 days prior to enrolment. Prior infection was determined when previous infections were reported in the questionnaire or when antibodies (NP antibodies during omicron period) were detected in the dried bloodspot at enrolment indicating a previous infection. Prior immunity was defined by the presence of antibodies at enrolment, being vaccinated before enrolment, and/or self-reported prior enrolment infection.

### Statistical analysis

Our population for analysis included all secondary household cases from the three cohorts. Index cases were excluded, because index case ascertainment strategies differed between cohorts, because of testing availability, and are inherently incomplete, which may select for more severe cases. We grouped cases by variant dominant period. Secondary cases from the RECOVER and CoKids cohorts were therefore assigned to the wild-type/Alpha variant, whereas cases from the VERDI-RECOVER cohort were assigned to the Omicron BA.1 and BA.2 period (see supplementary Figure 1 for variant prevalence over time from national surveillance^11^). Population demographic and vaccination characteristics were compared by variant period using proportions and medians with interquartile ranges (IQR).

We compared the symptoms and severity of secondary case episodes in the wild-type/Alpha variant dominant period to those in the Omicron dominant period stratified by age-category: child (age 0-11), adolescent: (age 12-17), and adult (age above 17) and visualized using bar-charts, heat plots, and spline charts. Missing diaries and severity scores were imputed using the symptoms and severity of the day before and after the missing value. Cases that did not complete any diary were excluded. For each symptomatic episode we computed the maximum severity score, referring to the day with highest reported score during the episode, and the cumulative severity score, referring to the sum of severity scores during the entire episode. We used Chi-square test for symptom frequency and Mann-Whitney U test for symptom duration, number of symptoms, maximum and cumulative severity score. Next, we explored the association between variant and symptoms using binomial and Gaussian multivariable regression models. Odds ratios (ORs) were computed for each respiratory and systemic symptom to quantify the association between variant and symptom frequency, adjusted for age, gender and prior immunity. Prior immunity was based on vaccination status, serology at baseline and/or a reported previous positive PCR or antigen test. For unvaccinated secondary cases with missing serology at baseline and no prior positive test, we assumed no prior immunity. In sensitivity analyses we repeated the multivariate analyses assuming the 18 persons with unknown baseline serology had a prior infection.

The multivariate analysis was repeated to assess the effect of vaccination status (primary series versus booster) on symptom burden and the symptom duration and severity. Since the distributions of diseases severity and duration were skewed, the data was log-transformed before estimating the mean difference in diseases severity and duration between variants. This analysis was restricted to adults during the Omicron dominant period, as this was the only age-group who were eligible for booster vaccination at the time.

Statistical analyses were performed with R version 4.0.3 (R Core Team, Vienna, Austria). T-test and Chi-square and multivariate logistic regression were used for statistical analysis with Holm-Bonferroni correction (p < 0.05).

## Results

In total, 355 secondary household cases were detected across the three cohorts. 9 cases were excluded because of missing/incomplete diaries (see details on data completeness in Supplementary Table 2) or an infection with the Delta variant (1 case). For the remaining participants, data completeness was > 99% for diaries and severity scores, 80-87% for NTS samples and 66-96% for DBS. Of 346 cases included in analysis, 216 occurred during the wild-type/Alpha period and 130 during the Omicron period. Age varied between 0 and 78 years and the majority were adults (57.8%, Table 1). The proportion of children < 12 years of age was slightly higher during the Omicron period (38.5% vs 26.4%). During the wild-type/Alpha period, 6 cases (2.8%) had a prior SARS-CoV-2 infection and no cases had received any vaccination. In contrast, during the Omicron period, 37.7% had a prior SARS-CoV-2 infection and 63 (96.9%) of adults had received at least the primary series and 45 (71.4%) a booster vaccination. Of adolescents, 14 (93.3%) had received primary vaccination, whereas children < 12 years were all unvaccinated. Of the individuals with available serology test results, 3.6% (6/165) had NP antibodies during wild-type/Alpha period and 35.7% (40/112) during the Omicron period.

**Table 1.** Baseline characteristics secondary household cases.

|  | Mean (SD) or No. (%) |  |  |  |  |  |
| --- | --- | --- | --- | --- | --- | --- |
|  | Wild-type/Alpha Period |  |  | Omicron Period |  |  |
|  | Adult (≥18 years)<br>(n=135) | Adolescent (12-17 years)<br>(n=24) | Child (<12 years)<br>(n=57) | Adult (≥18 years) (n=65) | Adolescent (12-17 years)<br>(n=15) | Child (<12 years)<br>(n=50) |
| Mean age (SD) | 42.1 (13.7) | 14.9 (1.6) | 6.1 (3.2) | 42.1 (5.0) | 13.9 (1.6) | 7.1 (3.1) |
| Female | 60 (44.4%) | 14 (58.3%) | 23 (40.4%) | 37 (56.9%) | 4 (26.7%) | 24 (48.0%) |
| Prior infection* |  |  |  |  |  |  |
| Yes | 2 (1.5%) | 0 (0.0%) | 4 (7.0%) | 29 (44.6%) | 5 (33.3%) | 15 (30.0%) |
| No | 133 (98.5%) | 24 (100.0%) | 53 (93.0%) | 33 (50.8%) | 9 (60.0%) | 27 (54.0%) |
| Unknown | 0 (0.0%) | 0 (0.0%) | 0 (0.0%) | 3 (4.6%) | 1 (6.7%) | 8 (16.0%) |
| Prior immunity# |  |  |  |  |  |  |
| Yes | 2 (1.5%) | 0 (0.0%) | 4 (7.0%) | 65 (100.0%) | 15 (100.0%) | 16 (32.0%) |
| No | 133 (98.5%) | 24 (100%) | 53 (93.0%) | 0 (0.0%) | 0 (0.0%) | 34 (68.0%) |
| Vaccination status |  |  |  |  |  |  |
| Primary plus booster series | 0 (0.0%) | 0 (0.0%) | 0 (0.0%) | 45 (69.2%) | 0 (0.0%) | 0 (0.0%) |
| Primary series | 0 (0.0%) | 0 (0.0%) | 0 (0.0%) | 18 (27.7%) | 14 (93.3%) | 0 (0.0%) |
| Incomplete primary series | 0 (0.0%) | 0 (0.0%) | 0 (0.0%) | 2 (3.1%) | 0 (0.0%) | 1 (2.0%) |
| No vaccination | 135 (100.0%) | 24 (100%) | 57 (100.0%) | 0 (0.0%) | 1 (6.7%) | 49 (98.0%) |

\*Prior infection is defined as having NP antibodies at enrolment and/or a reported previous infection in the baseline questionnaire.
#Prior immunity is defined as reported vaccinated before enrolment, having NP antibodies at enrolment, or a reported previous infection in the baseline questionnaire.

### Symptom burden during wild-type/Alpha and Omicron periods

Table 2 and Figures 1-3 show the SARS-CoV-2 symptom burden per period stratified by age-group. Infection remained asymptomatic in 13.0% versus 6.2% (p=0.045) of cases in the wild-type/Alpha versus Omicron period, less pauci-symptomatic and more symptomatic disease was observed in the Omicron period (63.4% versus 75.4%, p=0.021). Overall, the symptom burden was highest in adults during the wild-type/Alpha period and differences across age-groups largely disappeared during the Omicron period. There were diverging trends in symptom burden between children and adults. Among adults, maximum and cumulative severity scores tended to be lower during the Omicron period (p=0.06 and p=0.024). Loss of smell or taste was significantly less common during the Omicron period (p=0.001), whereas sore throat was more common (p=0.017; Figure 1-3). The number of adolescents included in analysis was low, but data suggest a higher proportion of symptomatic disease during the Omicron period (86.7% versus 58.3%, p=0.062) and a higher number of symptoms during the Omicron period (p=0.076). In particular cough, sore throat, cold shivers, and fever, while disease severity was not increased. In children < 12 years, the number of reported symptoms and the maximum severity score were significantly increased during the Omicron period (p≤0.011, Table 2 and Figure 1-3). In particular fatigue was more common during the Omicron period (p=0.004), and symptom duration was twice as long (3 days to 6 days, p=0.044).

**Table 2.** Symptom burden of SARS-CoV-2 infections Wild-type/Alpha versus Omicron variant per age group.

|  | No. (%) or median (IQR) |  |  |  |  |  |  |  |  |
| --- | --- | --- | --- | --- | --- | --- | --- | --- | --- |
|  | Adult >18 years |  |  | Adolescent 12-17 years |  |  | Child < 12 years |  |  |
|  | Wild-type/alpha period | Omicron Period |  | Wild-type/alpha period | Omicron Period |  | Wild-type/alpha period | Omicron Period |  |
|  | n=135 | n=65 | p-value<sup>§</sup> | n=24 | n=15 | p-value<sup>§</sup> | n=57 | n=50 | p-value<sup>§</sup> |
| Disease category |  |  | 0.046 |  |  | 0.139 |  |  | 0.034 |
| Symptomatic disease | 103 (76.3%) | 55 (84.6%) |  | 14 (58.3%) | 13 (86.7%) |  | 20 (35.1%) | 30 (60.0%) |  |
| Pauci-symptomatic | 20 (14.8%) | 10 (15.4%) |  | 7 (29.2%) | 2 (13.3%) |  | 24 (42.1%) | 12 (24.0%) |  |
| Asymptomatic | 12 (8.9%) | 0 (0.0%) |  | 3 (12.5%) | 0 (0.0%) |  | 13 (22.8%) | 8 (16.0%) |  |
| Days with symptoms | 13 (7.5-21) | 10 (6-18) | 0.095 | 7 (1-12) | 6 (4-8) | 0.7 | 3 (1-7) | 6 (4-11.5) | 0.044 |
| Number of symptoms | 6 (3-6) | 5 (4-7) | 0.3 | 2.5 (1-5.5) | 4 (3-6.5) | 0.076 | 2 (1-3) | 3 (1.5-5) | **0.004** |
| Maximum severity score<sup>§</sup> | 14 (10-18) | 11 (9.5-15) | 0.066 | 11 (7.5-19.5) | 10 (10-12) | 0.8 | 7 (4-11) | 10 (8.5-14) | **0.011** |
| Cumulative severity score<sup>§</sup> | 80.5 (55.5-128) | 54.0 (33.5-115) | 0.024 | 40.5 (22-128) | 34 (28-62) | >0.9 | 20.5 (14-49) | 40.5 (28-69.5) | 0.075 |

<sup>§</sup> calculated for cases with symptomatic disease
<sup>§</sup> Bold p-values indicate significant differences after correction for multiple testing.

**Figure 1.**
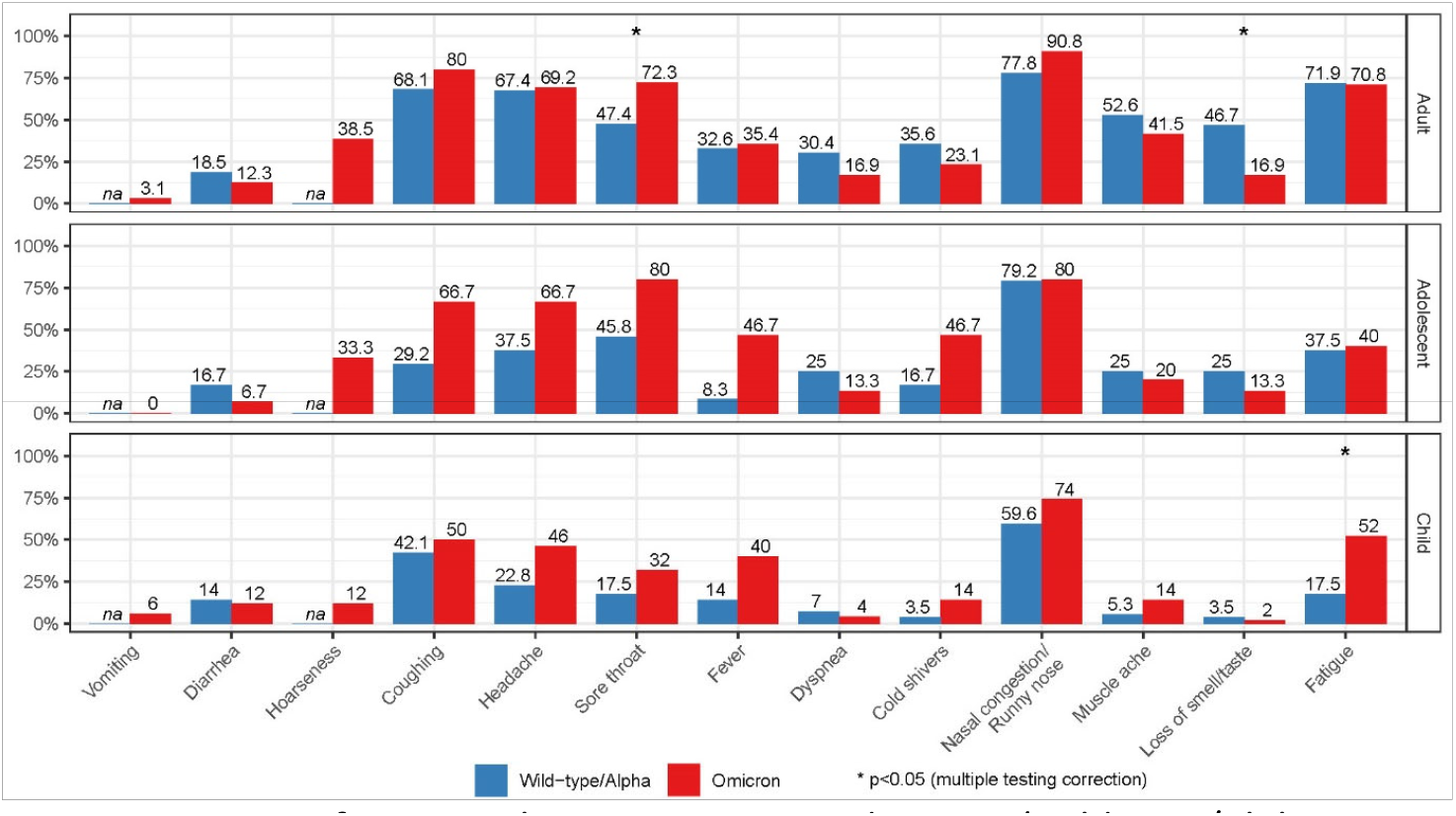
Symptom frequency by age-category and variant (Wild-type/Alpha versus Omicron)

**Figure 2.**
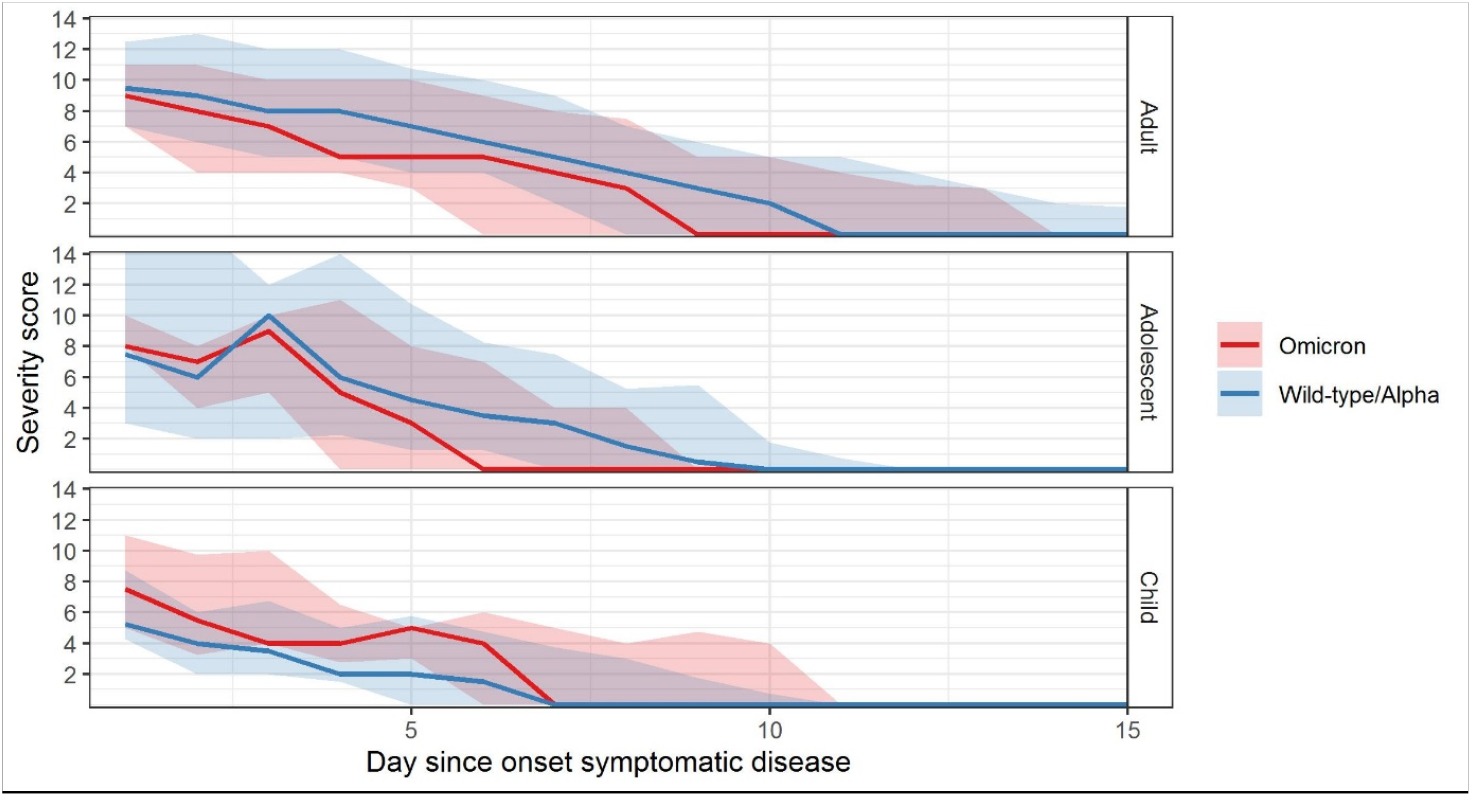
Symptom severity scores over time for symptomatic SARS-CoV-2 episodes by Wild-type/Alpha versus Omicron variant and age-group. Line indicate median and shade interquartile range

**Figure 3.**
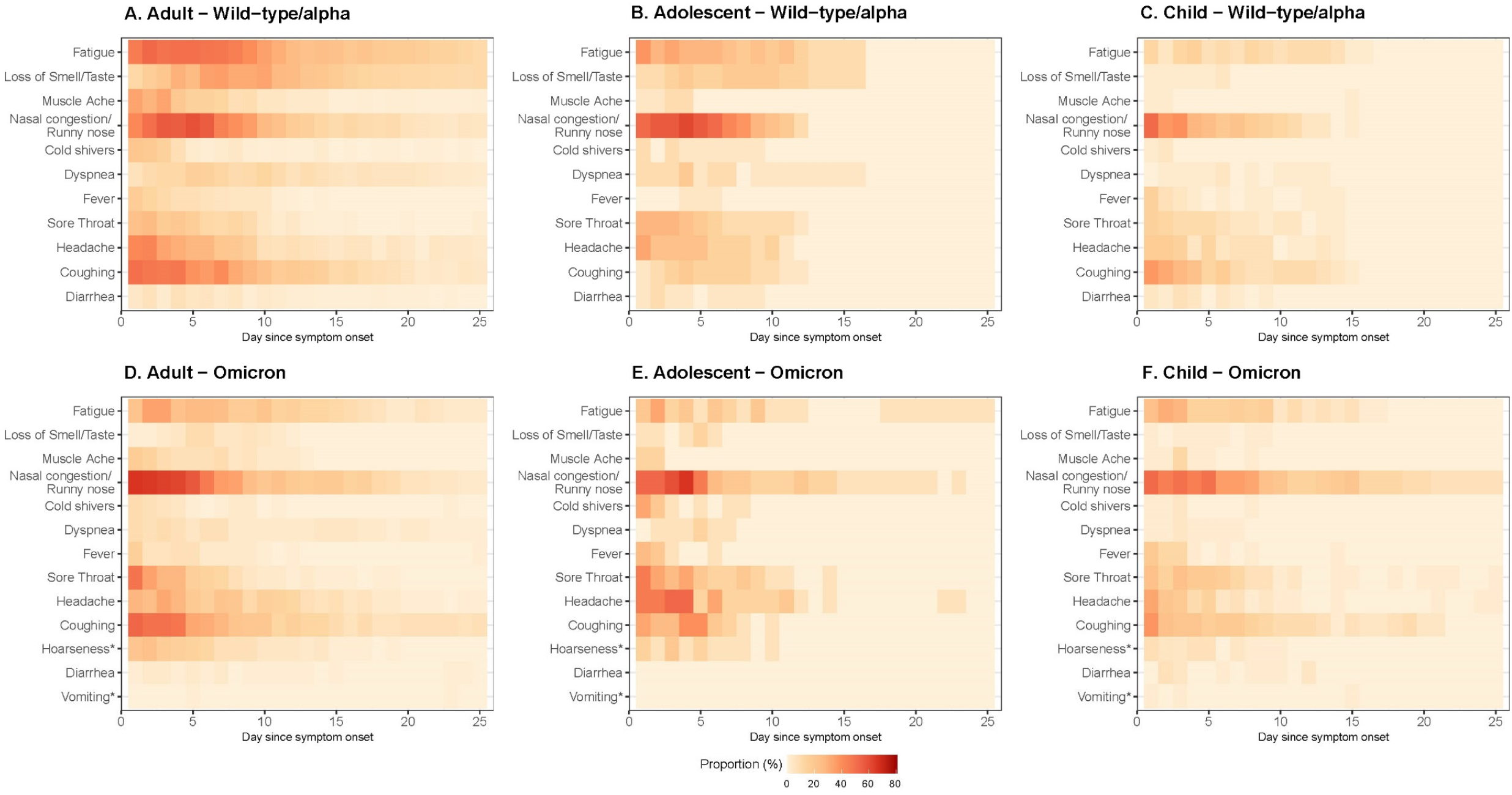
Symptom frequency per day since onset, age-group and variant. Includes all infected subjects (including asymptomatic cases). * indicate symptoms exclusively asked during omicron period.

After adjustment for age, gender, and prior immunity, the Omicron variant was associated with lower odds of loss of smell or taste (OR:0.14; 95%CI 0.03-0.50), whereas the Odds ratios for coughing (OR:1.85; 95%CI 0.92-3.78), fever (OR:2.23; 95%CI 1.03-4.81), sore throat (OR: 1.89; 95% CI 0.92-3.88), nasal congestion/runny nose (OR:1.97; 95%CI 0.89-4.59), and fatigue (OR:2.00; 95%CI 0.98-4.14) suggest and increase in these symptoms compared to the wild-type/Alpha variant (Table 3). Sensitivity analyses assuming the 18 persons with unknown NP antibodies at enrolment as infected, show equal trends in similar directions.

**Table 3.** Adjusted odds ratios for symptom frequency for Omicron versus Wild-type/alpha variant. No significant OR’s after correction for multiple testing.

| Symptom | OR (95% CI) | P-value<sup>§</sup> |
| --- | --- | --- |
| Muscle ache | 0.67 (0.25-1.67) | 0.4 |
| Coughing | 1.85 (0.92-3.78) | 0.085 |
| Fever | 2.23 (1.03-4.81) | 0.041 |
| Sore throat | 1.89 (0.92-3.88) | 0.082 |
| Nasal congestion/ Runny nose | 1.97 (0.89-4.59) | 0.103 |
| Dyspnea | 0.60 (0.16-1.89) | 0.4 |
| Headache | 1.39 (0.68-2.82) | 0.4 |
| Cold shivers | 1.00 (0.37-2.56) | 1.0 |
| Fatigue | 2.00 (0.98-4.14) | 0.058 |
| Diarrhea | 0.93 (0.32-2.50) | 0.9 |
| Loss of smell/taste | 0.14 (0.03-0.50) | 0.005 |
| An OR > 1 indicates higher odds during the Omicron period. |  |  |
| <sup>§</sup> Bold p-values indicate significant differences after correction for multiple testing. |  |  |

### Vaccination status and symptom burden in adults during the Omicron period

Comparing subjects who had received the primary series of SARS-CoV-2 vaccination to those who had also received a booster dose, no significant differences were observed for any symptom or symptom burden outcome measure (Table 4-5; Supplement Figure 3-5). Sensitivity analysis assuming three subjects with unknown NP antibodies at enrolment as prior immune, showed similar results.

**Table 4.** Symptom burden among SARS-CoV-2 infected Omicron adults stratified by vaccination status.

|  | No. (%) or median (IQR) |  |  |
| --- | --- | --- | --- |
|  | Primary series | Primary plus booster series |  |
|  | n=18 | n=45 |  |
|  |  |  | p-value |
| Disease severity |  |  | 0.5 |
| - Symptomatic | 16 (88.9%) | 37 (82.2%) |  |
| - Pauci-symptomatic | 2 (11.1%) | 8 (17.8%) |  |
| - Asymptomatic | 0 (0%) | 0 (0%) |  |
|  |  |  | p-value |
| Days with symptom | 10 (6.5-15) | 11 (6-18) | 0.4 |
| Number of symptoms | 5 (4-7) | 5 (4-6) | 0.8 |
| Maximum severity score<sup>§</sup> | 10 (9-12) | 12 (10-15) | 0.088 |
| Cumulative severity score<sup>§</sup> | 47 (28.5-75.5) | 64 (34-136) | 0.24 |

**Table 5.** Adjusted odds ratios for symptom frequency and severity in primary versus primary plus booster vaccinated subjects among Omicron infected adults.

| Symptom | OR (95% CI) | P-value |
| --- | --- | --- |
| Hoarseness | 0.56 (0.15-1.95) | 0.4 |
| Nasal congestion/ Runny nose | 0.65 (0.03-5.83) | 0.7 |
| Muscle ache | 1.21 (0.38-4) | 0.7 |
| Coughing | 1.18 (0.27-4.58) | 0.8 |
| Fever | 1.44 (0.43-5.35) | 0.6 |
| Sore throat | 0.38 (0.07-1.45) | 0.19 |
| Dyspnea | 0.86 (0.19-4.55) | 0.8 |
| Headache | 0.62 (0.14-2.32) | 0.6 |
| Cold shivers | 2.22 (0.45-14.09) | 0.4 |
| Fatigue | 0.67 (0.15-2.56) | 0.6 |
| Diarrhea | 1.53 (0.28-12.31) | 0.6 |
| Loss of smell/taste | 5.89 (0.92-117.48) | 0.12 |
| Vomiting | ∞ (0-∞) | |
| Duration and severity | Absolute difference estimate (95% CI) | p-value |
| Days with symptom | 0.11 (-0.24-0.64) | 0.6 |
| Number of symptoms | 0.07 (-1.04-1.18) | 0.9 |
| Maximum severity score<sup>§</sup> | 0.16 (-0.04-0.42) | 0.13 |
| Cumulative severity score<sup>§</sup> | 25.55 (-20.78-71.87) | 0.3 |
| An OR/estimates > 1 indicates higher odds/estimates for persons with booster vaccination. |  |  |
| <sup>§</sup> calculated for cases with symptomatic disease. |  |  |

## Discussion

Our analysis on symptom data of secondary household cases of SARS-CoV2 in a non-hospitalized general community, detected by comprehensive screening, provides a detailed comparison of SARS-CoV2 disease profiles in the community during two periods. During the wild-type/Alpha variant period, the population immunity was low, while in the (early) Omicron period, when BA.1 and BA.2 subvariants were dominant, a large proportion of the adult and adolescent population had been vaccinated, but not children < 12 years. Interestingly, the symptom burden in children was higher during the Omicron period compared to the wild-type/Alpha period. In adults, there was a reduction in the number and duration of symptoms present during the Omicron period when nearly all of them had evidence of prior immunity from vaccination or previous infection. Trends in adolescents (93.3% vaccinated during Omicron period) were less clear, but overall numbers of infections in this age-group were low. Adjusted for age, gender and prior immunity Omicron was associated with lower odds for loss of smell or taste (Odds Ratio [OR]: 0.14; 95%CI 0.03-0.50), and higher, but non-significant odds for upper respiratory symptoms, fever and fatigue (ORs varying between 1.85-2.23). No significant differences in symptoms were observed between primary versus primary plus booster vaccinated adults during the Omicron period.

There is a general consensus that the Omicron BA.1 and BA.2 variant and its descendants cause less severe disease compared to previous variants of SARS-CoV-2^12,13^. This notion is largely based on the lower risk of SARS-CoV-2 hospitalization and in particular ICU admission, both in vaccinated and unvaccinated subjects, observed during the Omicron dominant period^7^. Our results demonstrate that the average symptom burden of SARS-CoV-2 respiratory illness in the community setting has not decreased substantially, and was even increased in (largely non-immune) children when compared to earlier variants. Symptoms of upper respiratory illness were more common during the Omicron period, while loss of smell and taste, a typical symptom of earlier variants, was infrequent. In adults and adolescents, there was also a nearly 50% reduction in dyspnea, but this was non-significant because of small sample size. This shift in disease symptoms for Omicron has been described by others^14^, and may be explained by the altered replication and cellular tropism of the Omicron variant in different compartments of the respiratory tract compared to earlier variants^14^. Several studies have described reduced replication of Omicron variants in lung parenchyma, but increased replication with higher viral loads in bronchial and nasopharyngeal mucosae.^15,16^ This may lead to more mucosal damage and inflammation, predominantly in the upper respiratory tract, which is then reflected in the symptom burden. This is particularly evident in subjects without prior immunity, i.e. unvaccinated children < 12 years of age in our study.

We found no difference in symptomatology between primary series vaccinated and primary plus booster series vaccinated cases during the omicron dominant period, but our sample size was small. A study on breakthrough infections among vaccinated health care workers showed that 11% of the primary series vaccinated participants were asymptomatic compared to 16% of the primary plus booster series vaccinated participants^17^, suggesting a small effect of boosters on symptom burden..

The interpretation of our results has some limitations. First, it is possible that coinfections with other pathogens influence disease severity and coinfection rates may vary over time. Based on National Virological Surveillance data from the Netherlands most respiratory viruses circulated at lower rates during the Omicron period compared to wild-type/Alpha period^18,19^. This could, therefore, not explain the higher symptom burden in children. Second, although similar protocol were used, the samples from the cohorts were tested at different laboratories (see Supplement Table 1), this may limit comparability to some extent. Third, we based prior infection status on the results of serology testing at enrolment and on self-reported history of positive PCR or antigen tests. Availability of PCR testing was limited in the first year of the pandemic and antibodies from prior infection may have been undetectable at the time of enrolment due to waning^20^. In our cohort, 44.6% of the adults had evidence of prior infection during the Omicron BA.1 and BA.2 period, but this is likely an underestimate. Lastly, the low number of adolescent participants yielded in insufficient power to obtain precise estimates for this age-group.

## Conclusion

In children, the Omicron variant causes higher symptom burden compared to the wild-type/Alpha. Adults experienced a lower symptom burden possibly due to prior vaccination. A shift in most frequently reported symptoms occurred with a marked reduction in loss of smell or taste during the Omicron period. An additional effect of booster vaccination on symptom severity in infected adults compared to primary series only, could not be demonstrated.

## Data Availability

All data produced in the present study are available upon reasonable request to the authors.

## Acknowledgements

We would like to thank all participants of the studies. In addition, we would like to thank Anne van der Linden for support with testing of the DBS specimens.

This work forms part of RECOVER (Rapid European COVID-19 Emergency Response research) and VERDI (SARS-coV2 variants Evaluation in pRegnancy and paeDIatrics cohorts). RECOVER (101003589) is funded by the EU Horizon 2020 research and innovation program. VERDI project (101045989) is funded by the European Union. Views and opinions expressed are however those of the author(s)only and do not necessarily reflect those of the European Union or the European Health and Digital Executive Agency. Neither the European Union nor the granting authority can be held responsible for them. In addition, part of the work is funded by ZonMw.

## References

1. World Health Organization. Tracking SARS-CoV-2 variants. World Health Organization. Published 2021. Accessed July 17, 2022. https://www.who.int/activities/tracking-SARS-CoV-2-variants

2. Fan Y, Li X, Zhang L, Wan S, Zhang L, Zhou F. SARS-CoV-2 Omicron variant: recent progress and future perspectives. Signal Transduct Target Ther. 2022;7(1):141. doi:10.1038/S41392-022-00997-X

3. Gupta R. SARS-CoV-2 Omicron spike mediated immune escape and tropism shift. Res Sq. Published online January 17, 2022. doi:10.21203/RS.3.RS-1191837/V1

4. Harvey WT, Carabelli AM, Jackson B, et al. SARS-CoV-2 variants, spike mutations and immune escape. Nat Rev Microbiol 2021 197. 2021;19(7):409–424. doi:10.1038/s41579-021-00573-0

5. Miller NL, Clark T, Raman R, Sasisekharan R. Insights on the mutational landscape of the SARS-CoV-2 Omicron variant. bioRxiv. Published online December 10, 2021:2021.12.06.471499. doi:10.1101/2021.12.06.471499

6. Aiello TF, Puerta-Alcalde P, Chumbita M, et al. Infection with the Omicron variant of SARS-CoV-2 is associated with less severe disease in hospitalized patients with COVID-19. J Infect. 2022;0(0). doi:10.1016/j.jinf.2022.07.029

7. Nyberg T, Ferguson NM, Nash SG, et al. Comparative analysis of the risks of hospitalisation and death associated with SARS-CoV-2 omicron (B.1.1.529) and delta (B.1.617.2) variants in England: a cohort study. Lancet. 2022;399(10332):1303–1312. doi:10.1016/S0140-6736(22)00462-7

8. Whittaker R, Greve-Isdahl M, Bøås H, Suren P, Buanes EA, Veneti L. COVID-19 Hospitalization Among Children <18 Years by Variant Wave in Norway. Pediatrics. 2022;150(3):2022057564. doi:10.1542/peds.2022-057564

9. Verberk JDM, de Hoog MLA, Westerhof I, et al. Transmission of SARS-CoV-2 within households: a remote prospective cohort study in European countries. Eur J Epidemiol. 2022;37(5):549–561. doi:10.1007/s10654-022-00870-9

10. de Hoog M, Sluiter-Post J, Westerhof I, et al. Incidence rates and symptomatology of community infections with SARS-CoV-2 in children and parents: The CoKids longitudinal household study. medRxiv. Published online December 11, 2021:2021.12.10.21267600. doi:10.1101/2021.12.10.21267600

11. Rijksinstituut voor Volksgezondheid en Milieu. Varianten van het coronavirus SARS-CoV-2. Published 2022. Accessed May 19, 2022. https://www.rivm.nl/coronavirus-covid-19/virus/varianten

12. Wolter N, Jassat W, Walaza S, et al. Early assessment of the clinical severity of the SARS-CoV-2 omicron variant in South Africa: a data linkage study. Lancet (London, England). 2022;399(10323):437–446. doi:10.1016/S0140-6736(22)00017-4

13. Ulloa AC, Buchan SA, Daneman N, Brown KA. Estimates of SARS-CoV-2 Omicron Variant Severity in Ontario, Canada. JAMA. 2022;327(13). doi:10.1001/JAMA.2022.2274

14. Vihta KD, Pouwels KB, Peto TE, et al. Omicron-associated changes in SARS-CoV-2 symptoms in the United Kingdom. medRxiv. Published online May 18, 2022:2022.01.18.22269082. doi:10.1101/2022.01.18.22269082

15. Meng B, Abdullahi A, Ferreira IATM, et al. Altered TMPRSS2 usage by SARS-CoV-2 Omicron impacts infectivity and fusogenicity. Nat 2022 6037902. 2022;603(7902):706–714. doi:10.1038/s41586-022-04474-x

16. Hui KPY, Ho JCW, Cheung M chun, et al. SARS-CoV-2 Omicron variant replication in human bronchus and lung ex vivo. Nat 2022 6037902. 2022;603(7902):715–720. doi:10.1038/s41586-022-04479-6

17. Robilotti E V, Whiting K, Lucca A, et al. Effectiveness of mRNA booster vaccine among health Care workers in New York City during the omicron surge, December 2021-January 2022. Clin Microbiol Infect. Published online August 3, 2022. doi:10.1016/J.CMI.2022.07.017

18. Nivel. Actuele weekcijfers aandoeningen – Surveillance. Published 2021. Accessed June 6, 2022. https://www.nivel.nl/nl/nivel-zorgregistraties-eerste-lijn/actuele-weekcijfers-aandoeningen-surveillance

19. Rijksinstituut voor Volksgezondheid en Milieu. Virologische weekstaten | RIVM. Rijksinstituut voor Volksgezondheid en Milieu. Published 2022. Accessed July 18, 2022. https://www.rivm.nl/surveillance-van-infectieziekten/virologische-weekstaten

20. Liu W, Russell RM, Bibollet-Ruche F, et al. Predictors of Nonseroconversion after SARS-CoV-2 Infection. Emerg Infect Dis. 2021;27(9):2454–2458. doi:10.3201/eid2709.211042

