## Supplement for "Symptom presentation among SARS-CoV-2 positive cases and the impact of COVID-19 vaccination; three prospective household cohorts"


##### Authors

Ilse Westerhof[1], Marieke de Hoog[1], Margareta Ieven[2], Christine Lammens[2], Janko van Beek[3], Ganna Rozhnova[1,4], Dirk Eggink[5], Sjoerd Euser[6], Joanne Wildenbeest[7], Liesbeth Duijts[8], Marlies van Houten[9,10], Herman Goossens[2], Carlo Giaquinto[11], Patricia Bruijning-Verhagen[1]

1. Julius Centre for Health Sciences and Primary Care, Department of Epidemiology, University Medical Centre Utrecht, Utrecht, The Netherlands
2. Laboratory of Medical Microbiology, Vaccine & Infectious Disease Institute, University of Antwerp, Antwerp, Belgium
3. Department of Viroscience, Erasmus University Medical Center Rotterdam, Rotterdam, The Netherlands
4. BioISI—Biosystems & Integrative Sciences Institute, Faculdade de Ciências, Universidade de Lisboa, Lisboa, Portugal
5. Centre for Infectious Disease Control, WHO COVID-19 Reference Laboratory, National Institute for Public Health and the Environment (RIVM), Bilthoven, The Netherlands.
6. Streeklaboratorium voor de Volksgezondheid Kennemerland, Department of Epidemiology, Kennemerland, The Netherlands ·
7. Department of Pediatric Infectious Diseases and Immunology, Wilhelmina Children's Hospital University Medical Center Utrecht, Utrecht, The Netherlands
8. Erasmus MC - Sophia Children's Hospital, Erasmus University Medical Center, Department of Pediatrics, Rotterdam, The Netherlands
9. Spaarne Gasthuis Academy, Spaarne Gasthuis, Hoofddorp, The Netherlands
10. Spaarne Gasthuis, department of Pediatrics, Haarlem and Hoofddorp, The Netherlands
11. Department of Women and Child Health, University of Padova, Padova, Italy

##### Corresponding Author

Ilse Westerhof (E:)
Julius Centre for Health Sciences and Primary Care, Department of Epidemiology, University Medical Centre Utrecht, Utrecht, The Netherlands, PO Box 85500, 3508 GA Utrecht

#### Supplement 1 Laboratory analyses VERDI-RECOVER household study

Nasopharyngeal swabs were stored immediately at -20°C after collection. Upon arrival in the Central lab, samples were thawed and aliquoted. One aliquot was used for further analysis. SARS-CoV-2 RNA extraction was performed using the MagMAXTMViral/Pathogen II Nucleic Acid Isolation Kit (Thermofisher Scientific) on a KingFisher™ Flex Purification System (Thermofisher Scientific, Waltham, MA, USA) with input and elution volumes of 200 µL and 50 µL, respectively.

RT-qPCR detection of the ORF1ab-, N- and S-genes was performed using the TaqPath™ COVID-19 RT-PCR kit on a QUANTSTUDIO™5 (Thermofisher Scientific). Data analysis was performed using FASTFINDER ANALYSIS software (v4.3.3, UgenTec, Hasselt, Belgium). Specimens with a cycle threshold (Cp-/Ct-values) less than or equal to 40 were defined as SARS-CoV-2 positive.

Dried blood spot specimens collected in the Omicron period were tested by multiplex protein microarray for antibodies targeting SARS-CoV-2 spike ectodomain and nucleoprotein as described previously 1. The SARS-CoV nucleoprotein was used to detect antibodies targeting the SARS-CoV-2 nucleoprotein due to antigen availability. We used a cohort of DBS specimens obtained from 61 SARS-CoV-2 PCR confirmed individuals (collected 30-92 days post symptom onset) and 31 serology negative baseline specimens collected in the RECOVER study for validation of a new batch of the SARS-CoV-2 spike ectodomain and SARS-CoV nucleoprotein antigens. A fluorescence cut off of 7000 and 900 for respectively the SARS-CoV-2 spike ectodomain and SARS-CoV nucleoprotein were chosen to optimize background fluorescence reactivity and sensitivity of the assay. The sensitivity and specificity using these cut offs were 0.92 and 0.97 for spike ectodomain and 0.93 and 0.97 for nucleoprotein.

Supplement Table 1 Overview study design **SARS-CoV-2 household studies**

| | RECOVER-household | | CoKids household study | | VERDI-RECOVER household study | |
|---|---|---|---|---|---|---|
| Inclusion criteria household composition | At least 3 persons in household, any ages | | At least 3 persons in household and at least one child < 18 years of age | | At least 3 persons in household and at least one child < 18 years of age | |
| Inclusion criteria SARS-CoV-2 infection | Household index case with positive PCR test result from a municipal or healthcare testing facility and enrolment within 48 hours | | Household participating in prospective Cokids follow-up and positive Index case on any of the household asymptomatic or symptomatic PCR screening tests in previous 48 hrs | | Household index case with positive PCR test result from a municipal or healthcare testing facility, or a positive antigen (self-test and enrolment within 48 hours | |
| Diary symptoms | ✓ | Muscle ache | ✓ | Muscle ache | ✓ | Muscle ache |
| | ✓ | Coughing | ✓ | Coughing | ✓ | Coughing |
| | ✓ | Fever | ✓ | Fever | ✓ | Fever |
| | ✓ | Sore throat | ✓ | Sore throat | ✓ | Sore throat |
| | ✓ | Nasal congestion/ Runny nose | ✓ | Nasal congestion/ Runny nose | ✓ | Nasal congestion/ Runny nose |
| | ✓ | Dyspnea | ✓ | Dyspnea | ✓ | Dyspnea |
| | ✓ | Headache | ✓ | Headache | ✓ | Headache |
| | ✓ | Cold shivers | ✓ | Cold shivers | ✓ | Cold shivers |
| | ✓ | Fatigue | ✓ | Fatigue | ✓ | Fatigue |
| | ✓ | Diarrhea | ✓ | Diarrhea | ✓ | Diarrhea |
| | ✓ | Loss of smell/taste | ✓ | Loss of smell/taste | ✓ | Loss of smell/taste |
| | - | Vomiting | ✓ | Vomiting | ✓ | Vomiting |
| | - | Hoarsness | - | Hoarsness | ✓ | Hoarsness (added January 17, 2022) |
| Sample collection | | | | | | |
| NTS | ✓ | Enrolment | ✓ | Enrolment | ✓ | Enrolment |
| | ✓ | Onset Symptomatic disease | ✓ | Onset Symptomatic disease | ✓ | Onset Symptomatic disease |
| Dry blood spot | ✓ | Enrolment | ✓ | Enrolment | ✓ | Enrolment |
| | ✓ | 10 days after last diary | ✓ | 10 days after last diary | ✓ | 10 days after last diary |
| Laboratory | | | | | | |
| NTS | Laboratory of Antwerp University, Antwerp, Belgium | | Laboratory of Streeklab Haarlem, the Netherlands | | Laboratory of Antwerp University, Antwerp, Belgium | |
| DBS | Erasmus University Medical Center Rotterdam, Rotterdam, The Netherlands | | WHO COVID-19 Reference Laboratory of National Institute for Public Health and the Environment (RIVM), Bilthoven, The Netherlands. | | Erasmus University Medical Center Rotterdam, Rotterdam, The Netherlands | |

Supplement Table 2 data completeness for 346 secondary household cases included in analysis.

| | **RECOVER n=170** | **CoKids n=46** | **VERDI-RECOVER n=130** |
|---|---|---|---|
| Diaries | | | |
| Persons with any missing | 2 (<1.0%) | 1 (2.2%) | 1 (<1.0%) |
| Number of missing dairies | 4 (<0.01%) | 2 (<0.01%) | 6 (<0.01%) |
| Severity scores | | | |
| Persons with missing | 4 (<1.0%) | 1 (2.2%) | 1 (<1.0%) |
| Number of missing severity scores | 8 (<0.01%) | 2 (<0.01%) | 5 (<0.01%) |
| Samples completeness | | | |
| NTS enrolment | 138 (81.2%) | 40 (87.0%) | 104 (80.0%) |
| DBS enrolment | 44 (95.7%) | 121 (71.2%) | 112 (86.2%) |
| DBS end of study | 35 (76.1%) | 146 (85.9%) | 85 (65.4%) |

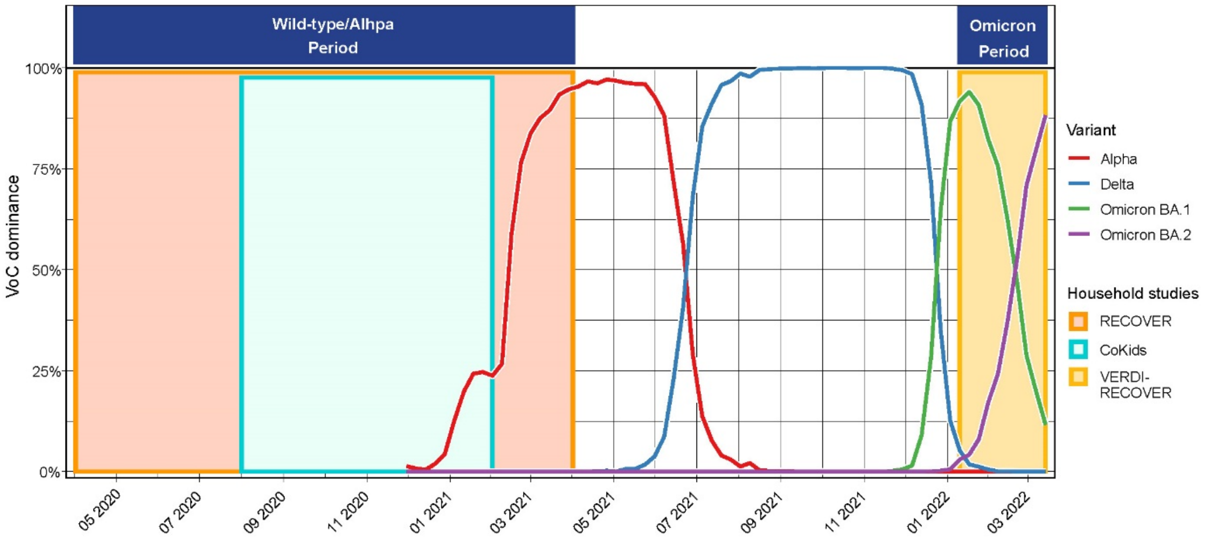


Supplement Figure 1 SARS-CoV-2 variants dominance in the Netherlands and study periods. Figure adapted from Dutch national surveillance[2].

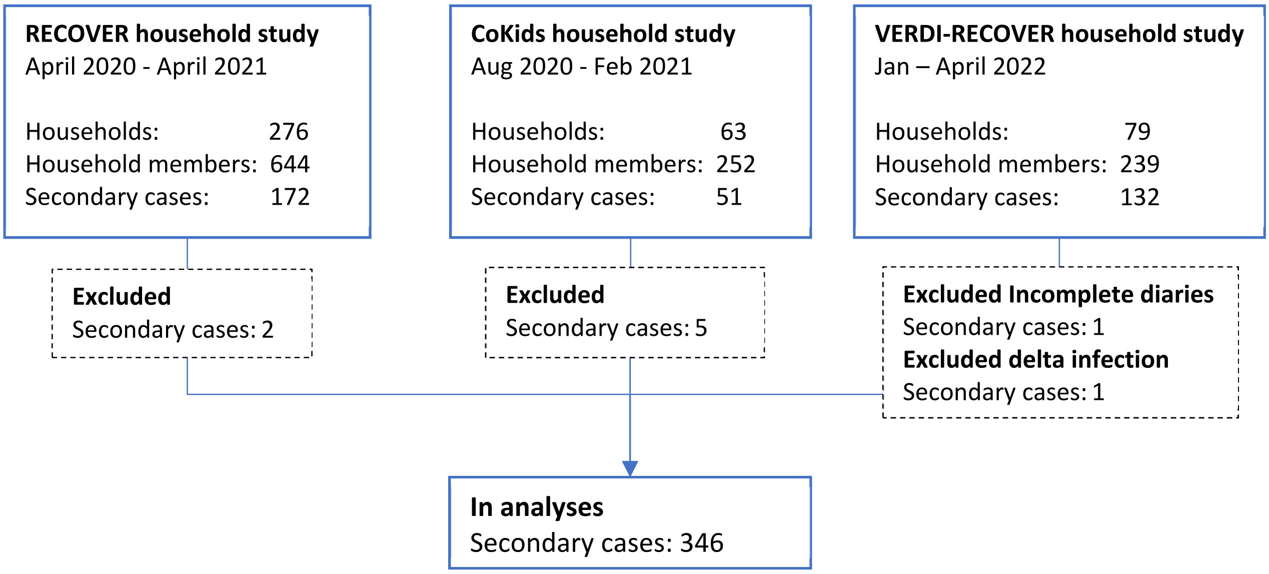


Supplement Figure 2 Flowchart of household studies. Secondary cases with NTS RT-PCR positive result at enrolment or at onset symptomatic disease. 98 secondary cases were excluded because of not completing any diary. One secondary case was excluded because of an infection with the Delta variant. A total of 346 were included in the analyses.

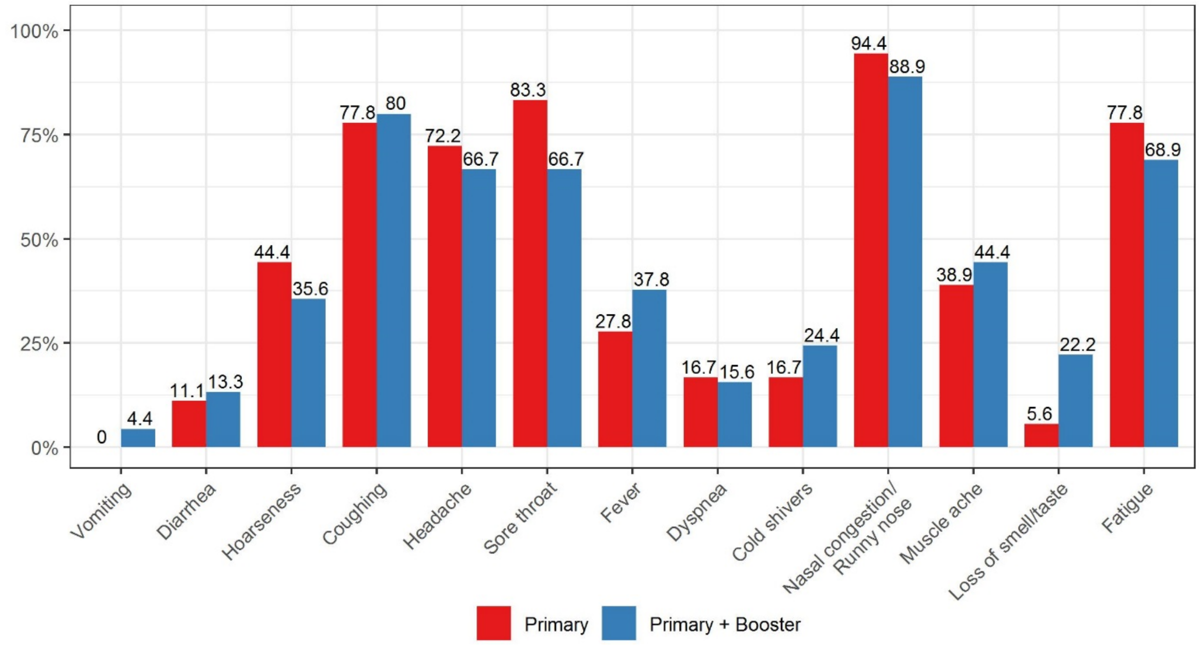


Supplement Figure 3 Symptomatology of SARS-CoV-2 infected adults during the omicron period by vaccination status.

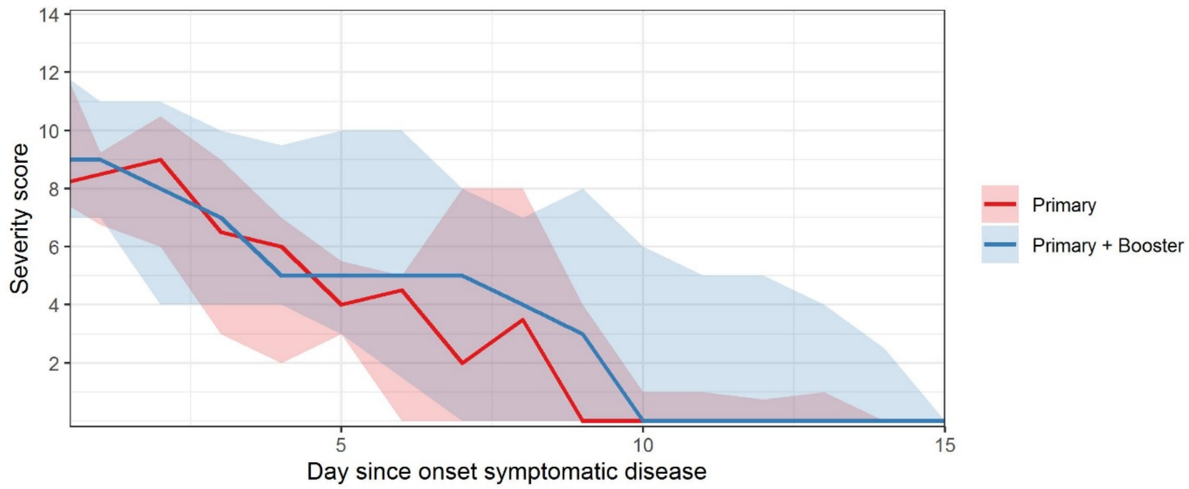


Supplement Figure 4 Severity scores over time for symptomatic SARS-CoV-2 episodes in adults by vaccination status. Line indicate median and shade interquartile range.

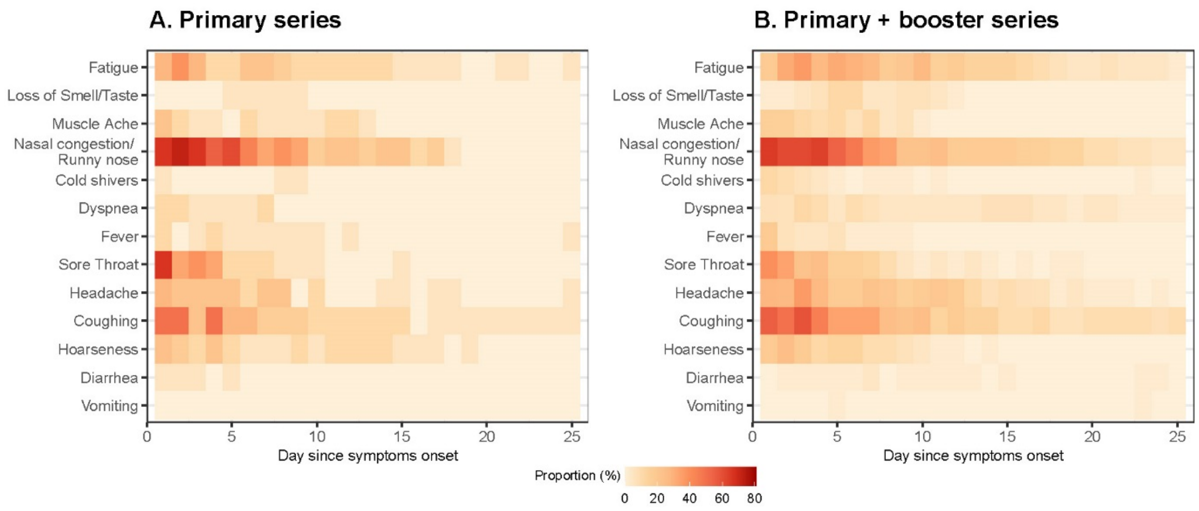


Supplement Figure 5 Symptom frequency by day since onset among primary or primary plus booster vaccinated adults with symptomatic SARS-CoV-2 infection during the Omicron dominant period.

### REFERENCES


1. Verberk JDM, de Hoog MLA, Westerhof I, et al. Transmission of SARS-CoV-2 within households: a remote prospective cohort study in European countries. *Eur J Epidemiol*. 2022;37(5):549-561. doi:10.1007/s10654-022-00870-9

2. Rijksinstituut voor Volksgezondheid en Milieu. Varianten van het coronavirus SARS-CoV-2. Published 2022. Accessed May 19, 2022. https://www.rivm.nl/coronavirus-covid-19/virus/varianten
